# Nurse specialist led sleep pathway is clinically effective and cost effective compared to a pathway delivered by consultants

**DOI:** 10.1101/2025.06.03.25328720

**Authors:** Marcus Pittman, Lisa Ward, Subimol Devasia, Wendy Pratley, Maria Darda, Dawn Cooke, Samantha Bloxham

## Abstract

**Introduction:** Professionals other than Physicians are commonly employed to evaluate patients with possible sleep apnoeas worldwide, but less so in the UK. This study evaluates a novel Nurse Specialist led sleep diagnostic pathway.

**Methods:** A sample of patients treated in the novel, Nurse led, pathway were compared to a group treated in a more conventional, Consultant led, setting.

**Results:** The use of a Nurse led pathway significantly reduced time from referral to CPAP set up (56 days) compared to the Consultant led pathway (213.5 days) with comparable outcomes in terms of CPAP compliance and symptom improvement.

**Conclusion:** The use of a Nurse led sleep diagnostic pathway significantly reduces waiting times, whilst maintaining appropriate clinical standards.

## Introduction

Nurses and midwives are increasingly taking on more complex, autonomous and expert roles, commonly referred to as ‘advanced practice’; with numbers undertaking such roles continuing to rise, even in recent years.^1^ In the US, Nurse Practitioners and Physicians Assistants are widely employed in clinical roles in sleep clinics, and have been for some time.^2^ However, numbers of Physician Associates,^3^ and Nurse Practitioners^1^ employed in the UK health service are considerably lower than in the US.^4^ The fact that sleep clinics see a high volume of patients, often with the same diagnosis of obstructive sleep apnoea (OSA) is perhaps a situation that lends itself well to the use of professionals other than Consultant Physicians to carry out significant proportions of this work, and in addition allows assessment and treatment to be conducted in the same appointment.

The merger of two hospitals in the United Kingdom of a similar size, serving a similar demographic, but with very different sleep pathways, allowed a comparison to be conducted in two main areas; the use of Clinical Nurse Specialists (CNS) compared to the use of Consultant Physicians, and of the use of auto-adjusting positive airway pressure (APAP) devices compared to the use of fixed pressure continuous positive airway pressure (CPAP) machines. There are potential cost implications for both areas of comparison.

The pathway on the consultant site consisted of General Practitioner (GP) referral, followed by a home sleep study, followed by assessment in a consultant out-patient clinic, followed by (for those who required it) APAP set up in a group setting run by a Band 8B physiologist, supported by a Band 3 assistant. Routine follow up was timetabled for 3 months with a Band 3, but with the provision of a ‘walk-in’ service (provided by Band 3, 6, or 8B team members) for troubleshooting.

The novel pathway on the CNS site consisted of GP referral, followed by a home sleep study, followed by assessment by a Band 7 or Band 8A CNS with CPAP set up at the same appointment by a Band 4 or Band 5 Health Care Sleep Assistant. Follow up was timetabled based on individual patient need, with the provision of ad hoc telephone or face to face appointments if required for CPAP titration and/or troubleshooting.

## Methods

A retrospective study was conducted on patients set up with positive airway pressure (PAP) therapy on the two sites, sampled from August to December 2018; allowing a reasonable follow up period before the covid pandemic started. As the novel service on the CNS site involved a significant change in practice, thorough data on all patients were collected. A smaller sample of the more conventional consultant led site were collected to allow a comparison to be conducted. Data were collected on time from referral to CPAP set up, severity, patient outcomes, and approximate costs of the variable parts of the pathways. A total of 132 episodes were investigated.

Staff pay levels were calculated at 2024 pay scales, to bring the cost comparison up to date^5, 6^, and converted into hourly rates^7^.

## Results

All patients on the consultant led site saw a Consultant Physician at least once and were all given an auto-adjusting positive airway pressure (APAP) device. None of the patients on the CNS led site required a consultant appointment, and only 1 of the 90 patients was given an APAP device, the rest receiving a fixed pressure continuous positive airway pressure (CPAP) machine. Overall, these differences resulted in a lower set up cost for the novel CNS pathway.

Time from GP referral to PAP set up was significantly lower on the CNS site compared to the consultant site, 56 days versus 213.5 days respectively.

Interestingly severity was significantly higher in those set up with PAP on the CNS site compared to the consultant site, however a number of patients on the consultant site were given CPAP on the basis of compelling symptoms, even though they had essentially normal oximetry (oxygen desaturation index less than 5), which may explain at least part of this difference. A review of patients seen in the respective sleep clinics, but not started on PAP, indicated a significantly higher number of patients with mild OSA and minimal symptoms who were offered lifestyle advice and not started on PAP on the CNS site, compared to the consultant led site; in keeping with the current the then current National Institute for Health and Care Excellence guidance (2008)^8^ on the use of CPAP in the treatment of OSA.

Mean CPAP pressure used was significantly lower on the CNS site, which predominantly relied on fixed pressure machines, however as the compliance hours and change in Epworth sleepiness scale were not significantly different, this does not necessarily suggest under-treatment in this group. BMI was not different in the two patient groups, and thus does not explain the differences seen in severity or CPAP pressure.

**Table 1:** Mean (unless otherwise stated) figures comparing the patients, their outcomes (at approximately 3 months), and the staff costs of the two pathways.

|  | **Consultant led** | **CNS led** |
| --- | --- | --- |
| Number of Patients | 42 (12 Female) | 90 (30 Female) |
| Referral to PAP set up no days | 213.5 | 56.0 (p<0.01) |
| Desaturation Index at diagnosis | 20.5 | 39.1 (P<0.01) |
| Body Mass Index | 36.4 | 35.6 (NS) |
| Improvement in Epworth with PAP | 7.19 | 5.6 (NS) |
| Compliance Hours | 5.36 | 5.73 (NS) |
| Proportion Compliant | 61.9% | 67.6% |
| CPAP Pressure | 12.21 | 10.49 (p<0.01) |
| Number of patient visits | 5.83 | 4.84 |
| Estimated variable staff costs | £60.10 | £38.23 |

**Table 2:** Assessment and CPAP set up staff costs per patient (excluding home sleep study cost). NB in the CNS pathway, the assessment and set up appointments occur at the same visit.

|  | Consultant led | CNS led |
| --- | --- | --- |
| Assessment appointment | 27 minutes consultant time<br>£24.86 | 15 minutes CNS time<br>£6.79 |
| CPAP set up | 4 hours of 8B & B3 for 20 patients<br>£8.84 | 30 minutes Band 4 or 5 time<br>£7.39 |
| Total per patient | £33.70 | £14.18 |

**Table 3:** Stabilisation and follow up, first three months after CPAP set up; staff costs.

|  | Consultant led | CNS led |
| --- | --- | --- |
| Planned follow up appointment | 30 minutes of B3 time<br>£6.17 | 15 minutes of B7 time<br>£5.83 |
| Titration or Ad hoc appointments | 20 minutes of B8B, B6 or B3 time,<br>average 2.83 visits = £20.23 | 15 or 20 minutes of B5, B7, or B8A<br>time average 2.84 visits = £18.22 |
| Total | £26.40 | £24.05 |

## Discussion

The use of CNS significantly reduced waiting times for CPAP set up by expanding the number of new appointments available. Compliance hours and change in Epworth score were comparable between the two sites. The number of patient visits on the consultant led site was higher than the CNS led site, as on the CNS site the first appointment involved initial assessment and PAP set up at the same time, which also contributed to the lower referral to set up time.

The fact that some patients were diagnosed with OSA and set up with PAP therapy after a normal sleep study (albeit in the context of compelling symptoms) on the consultant site, and that more patients on the CNS site with mild OSA and minimal symptoms were given lifestyle advice and not started on PAP, suggests practice on the CNS site adhered more closely to the then current National Institute for Health and Care Excellence guidance (2008)^8^ on the use of CPAP in the treatment of OSA, which is also in keeping with the most recent guidance (2021)^9^, with further potential implications for the cost-effectiveness for this approach.

Overall, the novel CNS led sleep clinics investigated in this study increased capacity, and thereby shortened the time from referral to treatment, and also reduced staff costs compared to the traditional consultant led pathway, without negatively impacting patient outcomes. The use of autoset devices did not significantly influence compliance and sleepiness compared to fixed pressure machines in this study. Changes in the relative costs of APAP machines compared to fixed pressure CPAP devices may mean the current cost difference of this part of the pathway may be less important going forwards.

## Data Availability

All data produced in the present work are contained in the manuscript

